# Three separate spike antigen exposures by COVID-19 vaccination or SARS-CoV-2 infection elicit strong humoral immune responses in healthcare workers

**DOI:** 10.1101/2022.03.06.22271718

**Authors:** Thomas Theo Brehm, Felix Ullrich, Michelle Thompson, Julia Küchen, Dorothee Schwinge, Anthea Spier, Samuel Huber, Johannes K Knobloch, Martin Aepfelbacher, Marylyn M Addo, Ansgar W Lohse, Marc Lütgehetmann, Julian Schulze zur Wiesch

**Affiliations:** I. Department of Medicine, University Medical Center Hamburg-Eppendorf, Martinistraße 52, 20249 Hamburg, Germany; German Center for Infection Research (DZIF), Partner Site Hamburg-Lübeck-Borstel-Riems; Institute of Medical Microbiology, Virology and Hygiene, University Medical Center Hamburg-Eppendorf, Martinistraße 52, 20249 Hamburg, Germany; Institute for Infection Research and Vaccine Development, University Medical Center Hamburg-Eppendorf, Hamburg, Germany

**Author notes:** **Corresponding author:** Thomas Theo Brehm, I. Department of Medicine, University Medical Center Hamburg-Eppendorf, Martinistraße 52, 20246 Hamburg. contributed equally (Marc Lütgehetmann, Julian Schulze zur Wiesch).

**Keywords:** SARS-CoV-2, COVID-19, healthcare worker, vaccination, immunity, Germany

## Abstract

As part of our ongoing prospective seroprevalence study, we assessed the SARS-CoV-2 infection and COVID-19 vaccination-induced immunity of 697 hospital workers at the University Medical Center Hamburg-Eppendorf between January 17 and 31, 2022. The overall prevalence of anti-NC-SARS-CoV-2 antibodies indicating prior infection was 9.8% (n=68) and thus lower than the seroprevalence in the general population in Hamburg. At the current study visit, 99.3% (n=692) had received at least one vaccine dose and 93.1% (n=649) had received at least three vaccine doses. All vaccinated individuals had detectable anti-S1-RBD-SARS-CoV-2 antibodies (median AU/ml [IQR]: 13’891 [8’505 – 23’543]), indicating strong protection against severe COVID-19. In addition, we show that individuals who received three COVID-19 vaccine doses (median AU/ml [IQR]: 13’856 [8’635 – 22’705]), and those who resolved a prior SARS-CoV-2 infection and received two COVID-19 vaccine doses (median AU/ml [IQR] 13’409 [6’934 – 25’000]) exhibited the strongest humoral immune responses. The low prevalence of anti-NC-SARS-CoV-2 antibodies indicates persistent effectiveness of established infection control interventions in preventing nosocomial SARS-CoV-2 transmission with the delta and omicron viral variants as predominant strains. Our study further indicates that three exposures to the viral spike protein by either SARS-CoV-2 infection or COVID-19 vaccination are necessary to elicit particularly strong humoral immune responses, which supports current vaccination recommendations.

## Introduction

Since the onset of the coronavirus disease 2019 (COVID-19) pandemic, we have been conducting a prospective severe acute respiratory syndrome coronavirus type 2 (SARS-CoV-2) seroprevalence study among hospital workers at the University Medical Center Hamburg-Eppendorf. Previously, we have reported seroprevalence during the first months of the pandemic (Brehm et al., 2021a) and at the end of the third wave in May 2021 (Brehm et al., 2021b). At the latter time point, we could demonstrate a low SARS-CoV-2 infection rate of 4.7% and robust vaccine-induced humoral immune responses when most study participants had received primary COVID-19 vaccination with two vaccine doses. In the meantime, there was an unprecedented surge in SARS-CoV-2 infections in Germany in the fall of 2021, driven by seasonality, the emergence of the delta variant (B.1.617.2), and, more recently, the omicron variant (B.1.1.529) (Karim and Karim, 2021). In response, the administration of a third dose of an mRNA vaccine, also referred to as booster, was recommended in Germany in prioritized groups including healthcare workers in October 2021 (RKI 2021a) and for the general population in December 2021 (RKI 2021b) to address potential waning immunity over time and reduced effectiveness against the novel SARS-CoV-2 variants. As of January 2022, the population uptake of two doses of COVID-19 vaccines in Germany was 74.0%, and 53.1□% received an additional vaccine dose (RKI 2022). After adenovirus-vector vaccine-related thromboembolic issues were reported, the administration of AZD1222 (Vaxzevria®, AstraZeneca) was only continued in vaccinees older than 60 years in Germany in March 2021. With this policy change, many HCW in Germany received a heterologous primary vaccination with one dose of AZD1222 followed by either BNT162b2 (Comirnaty®, BioNTech/Pfizer) or mRNA-1273 (Spikevax®, Moderna) as the second dose. Previously, studies by us and others have suggested that this heterologous primary vaccination with AZD1222 followed by an mRNA vaccine may confer more potent humoral immune responses than a homologous vaccination regimen (Barros-Martins et al., 2021; Borobia et al., 2021; Brehm et al., 2021b; Pozzetto et al., 2021). Due to these findings and the advantage of a shorter time interval between vaccine doses, German national guidelines now recommend the administration of an mRNA vaccine for all vaccinees who had received one dose of AZD1222 regardless of age (RKI 2021c). Since only mRNA vaccines are currently recommended as third vaccine doses, many individuals with homologous primary vaccination with AZD1222 have now received heterologous boosting with either BNT162b2 or mRNA-1273. The immunogenicity of the various possible COVID-19 vaccine combinations in naïve and convalescent individuals has not been formally tested in controlled studies, and real-life observational studies comparing immunogenicity are scarce. To assess the SARS-CoV-2 infection and COVID-19 vaccination-induced immunity among employees of the University Medical Center Hamburg-Eppendorf, we performed another study visit between January 17 and 31, 2022, as part of our ongoing seroprevalence study.

## Materials and methods

### Study design

Hospital workers of the University Medical Center Hamburg-Eppendorf participating in our ongoing longitudinal study were invited to provide a serum sample between January 17 and 31, 2022. Prior SARS-CoV-2 infections and COVID-19 vaccination status were assessed using an online REDcap electronic data capture tool that had been specifically designed for the present study. The study protocol was reviewed and approved by the Ethics Committee of the Medical Council of Hamburg (PV 7298), and written informed consent was obtained by all study participants before recruitment. The two currently licensed mRNA vaccines, BNT162b2 and mRNA-1273, were subsequently subsumed to the single category “mRNA vaccines”.

### Serology

Antibodies against the receptor-binding domain (RBD) domain of the viral spike protein (S) were detected with the quantitative Elecsys anti-S1-RBD-SARS-CoV-2 assay (Roche, Mannheim, Germany; cut off 0.8 AU/ml), which has a reported sensitivity of 99.8% and a specificity of 100% (Roche 2022a). In addition, antibodies against the viral nucleocapsid protein (NC) were detected with the qualitative Elecsys anti-NC-SARS-CoV-2 Ig assay (Roche, Mannheim Germany; cut off ≥ 1 COI/ml), which has a reported sensitivity and specificity of 99.5% and 99.8%, respectively (Roche 2022b). Both electrochemiluminescence immunoassays (Cobas e411, Roche; Mannheim, Germany) use a double-antigen sandwich assay format that detects both IgA, IgM, and IgG. Samples with titers above 250 U/ml were manually diluted 1:100 in dilution buffer according to the manufacturer’s recommendations to increase the linear range to 25’000 U/ml.

### Statistical analyses

A two-tailed Mann Whitney U test was used to analyze median antibody titers between two subgroups, and Kruskal-Wallis was used to analyze median antibody titers between more than two subgroups. Linear regression was calculated among participants with three vaccine doses to predict the anti-S1-RBD-SARS-CoV-2 antibody titer from sex, age, different vaccination regimen, time since the booster vaccination, and presence of anti-NC-SARS-CoV-2 antibodies. Statistical analyses were performed using GraphPad Prism, version 9 for macOS (GraphPad Software, La Jolla, California, USA) and SPSS, version 26 for macOS (IBM Corporation, Armonk, NY, USA). P values less than 0.05 were considered statistically significant.

## Results

### Characterization of the study population

Between January 17 and 31, 2022, 697 hospital workers participated in the current study. This represents around 6% of all employees of the University Medical Center Hamburg-Eppendorf. Median age of the current study cohort age was 40 years (interquartile range [IQR] 31 – 50 years), 78.5% (n=547) of the participants were women (**Table 1**). Most study participants were healthcare workers directly involved in patient care at different departments of our tertiary care center: 37.4% were nurses (n=261), 20.1% physicians (n=140), 16.1% medical technicians (n=112), and 12.2% (n=85) had other professions The remaining 14.2% (n=99) were other employees not directly involved in patient care. Most study participants had received three vaccine doses (n=644; 92.4%), fewer were vaccinated once (n=5; 0.7%), twice (n=38; 5.5%) or four times (n=5; 0.7%). Five study participants (0.7%) had not been vaccinated yet.

**Table 1.** Characterization of the study population

| **Sex** |  |
| --- | --- |
| Male, n(%) | 143 (20.5) |
| Female, n(%) | 547 (78.5) |
| Diverse, n(%) | 7 (1.0) |
| **Age** |  |
| Median | 40 |
| IQR | 31 - 50 |
| **Profession** |  |
| Nurse, n(%) | 261 (37.4) |
| Physician, n(%) | 140 (20.1) |
| Medical Technician, n(%) | 112 (16.1) |
| Other healthcare worker, n(%) | 85 (12.2) |
| Non-healthcare worker, n(%) | 99 (14.2) |
| **COVID-19 vaccine doses** |  |
| 0, n(%) | 5 (0.7) |
| 1, n(%) | 5 (0.7) |
| *Ad26.COV2.S, n* | 3 |
| *mRNA, n* | 2 |
| 2, n(%) | 38 (5.5) |
| *AZD1222/AZD1222, n* | 1 |
| *AZD1222/mRNA, n* | 12 |
| *mRNA/mRNA, n* | 25 |
| 3, n(%) | 644 (92.4) |
| *AZD1222/AZD1222/AZD1222, n* | 2 |
| *AZD1222/AZD1222/mRNA, n* | 60 |
| *AZD1222/mRNA/mRNA, n* | 276 |
| *mRNA/mRNA/mRNA, n* | 303 |
| *N/A, n* | 3 |
| 4, n(%) | 5 (0.7) |
| *AZD1222/mRNA/mRNA/mRNA, n* | 1 |
| *mRNA/mRNA/mRNA/mRNA, n* | 4 |

### Serological results

All 692 study participants who had received at least one dose of a COVID-19 vaccine had detectable anti-S1-RBD-SARS-CoV-2 antibodies; the median titer was 13’891 AU/ml (IQR 8’505 – 23’528 AU/ml). The rate of individuals with anti-S1-RBD-SARS-CoV-2 antibody titers higher than 10’000 AU/ml increased from 8.3% (n=72) at the last study visit in May 2021 to 65.9% (n=459) at the current study visit (**Figure 1**). Anti-NC-SARS-CoV-2 antibodies indicating prior SARS-CoV-2 infection were now detected in 9.8% (n=68) individuals. Participants with and without detectable anti-NC-SARS-CoV-2 antibodies did not differ in median age and sex, and the rate of individuals with anti-NC-SARS-CoV-2 antibodies was similar among different professional groups (**Supplemental table 1**). Among study participants without prior SARS-CoV-2 infection reflected by the absence of anti-NC-SARS-CoV-2 antibodies, the median anti-S1-RBD-SARS-CoV-2 antibody titer significantly increased between those who received only one (median AU/ml [IQR]: 564 [24 – 3’070]) to those who received two (median AU/ml [IQR]: 1’663 [1’094 – 3’060]), three (median AU/ml [IQR]: 13’856 [8’635 – 22’705]) or four (median AU/ml [IQR]: 25’000 [25’000 – 25’000]) COVID-19 vaccine doses (p<0.0001) (**Figure 2**). Study participants who previously resolved a SARS-CoV-2 infection had significantly higher median anti-S1-RBD-SARS-CoV-2 antibody titers in the respective subgroups with two (median AU/ml [IQR] 13’409 [6’934 – 25’000] vs. 1’663 [1’094 – 3’060]; p<0.0001) and three (median AU/ml [IQR]: 24’393 [11’991 – 25’000] vs. 13’856 [8’635 – 22’705]; p<0.0001) COVID-19 vaccine doses. None of the five individuals who had received four vaccine doses had detectable anti-NC-SARS-CoV-2 antibodies therefore no statistical analysis was performed for this subgroup. We further compared anti-S1-RBD-SARS-CoV-2 antibody titers among anti-NC-SARS-CoV-2 antibody-negative study participants with three homologous versus heterologous COVID-19 vaccine doses (**Figure 3**). Those who received a heterologous vaccination regimen including both AZD1222 and mRNA vaccines (median AU/ml [IQR]: 15’105 [10’296 – 23’532]) had higher median antibody titers than those who received three mRNA vaccine doses (median AU/ml [IQR]: 11’973 [6’618 – 21’354]; p<0.0001). Among the HCW with heterologous COVID-19 vaccination regimen, no difference in immune response was observed between those with the vaccination regimen AZD1222/mRNA/mRNA (median AU/ml [IQR]: 14’980 [10’140 – 23’595]) compared to those with the vaccination regimen AZD1222/ AZD1222/mRNA (median AU/ml [IQR]: 17’171 [10’776 – 23’165]; p=3.6). Simple linear regression was carried out to investigate the relationship between anti-NC-SARS-CoV-2 antibodies and time since the booster vaccination among study participants with three vaccine doses (n=644). There was a significant negative linear relationship between the two (F(1, 642)=136.47, p<0.001, R^2^=0.18). The results of multiple linear regression analysis revealed that age, sex, vaccination regimen, time since the booster vaccination, and presence of anti-NC-SARS-CoV-2 antibodies significantly predicted the anti-S1-RBD-SARS-CoV-2 antibody titer (F(5, 638)=31.48, p<0.001, R^2^=0.20). Only time since the booster vaccination and presence of anti-NC-SARS-CoV-2 antibodies added statistically significantly to the prediction (p<0.001). Age, sex, and vaccination regimen were found not to be significant predictors of anti-S1-RBD-SARS-CoV-2 antibody titers (p>0.05). We further analyzed the kinetics of anti-S1-RBD-SARS-CoV-2 antibody titers among study participants without prior SARS-CoV-2 infection between the last study visit in May 2021 and the current study visit (**Supplemental Figure 1**). All but one of the 206 participants who had received one COVID-19 vaccine dose and who participated at the last study visit last year, and two additional COVID-19 vaccine doses up to the present study visit demonstrated an increase of the anti-S1-RBD-SARS-CoV-2 antibody titer (median AU/ml [IQR]: 60 [29 – 120] vs. 17’614 [11’538 – 25’000]; p<0.0001). Likewise, the vast majority (n=222, 88.4%) of the 251 study participants who were measured after two COVID-19 vaccine doses at the last study visit and now had three COVID-19 vaccine doses at the present study visit showed a further increase in humoral immune response. In comparison, 26 (10.4%) HCW had a decrease in antibody titers and three had persistently high antibody titers at the highest measurable concentration (median AU/ml [IQR]: 1’622 [692 – 5’039] vs. 11’780 [6’725 – 19’497]; p<0.0001).

**Figure 1.**
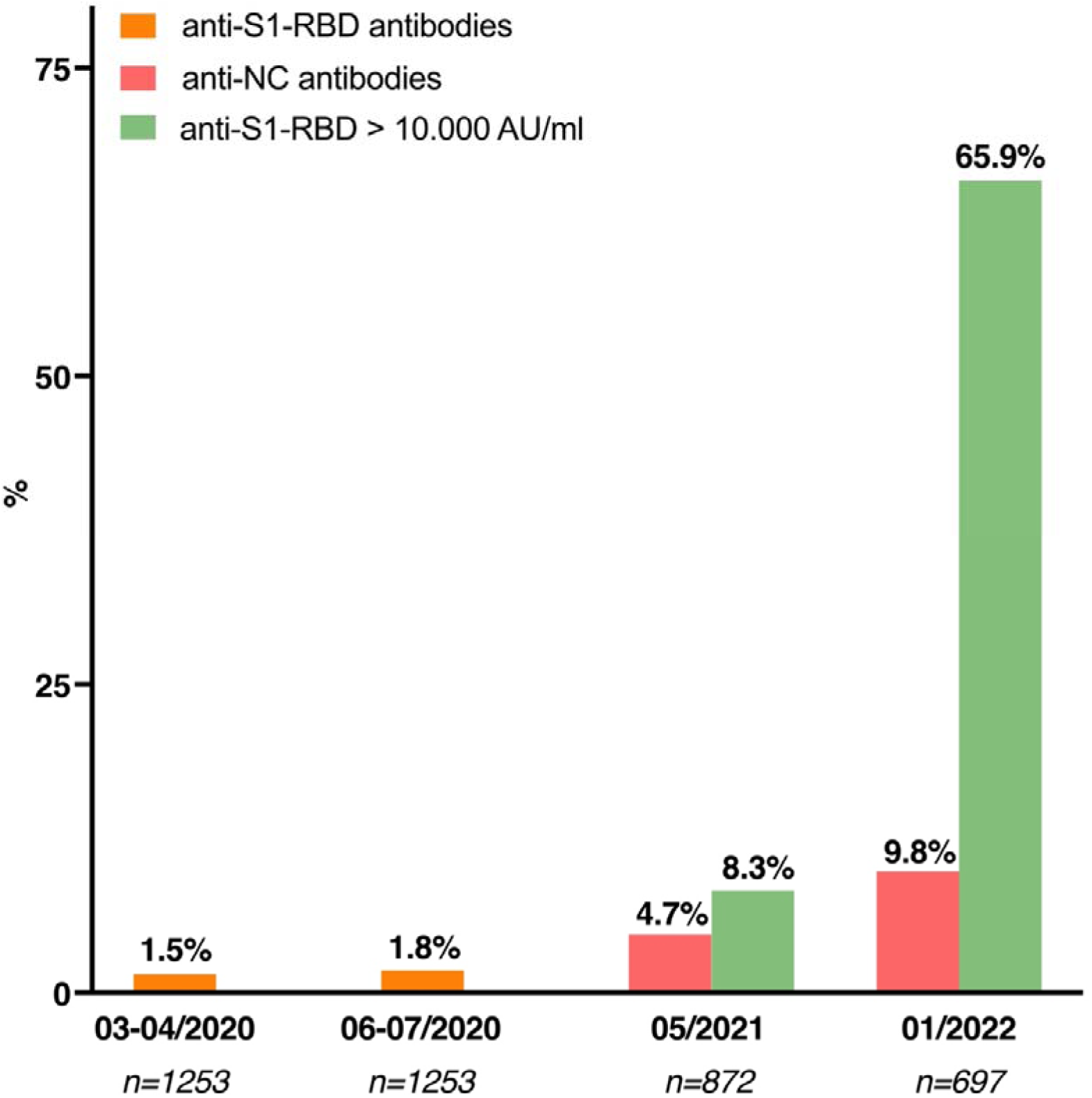
Results of our ongoing SARS-CoV-2 seroprevalence studies throughout the COVID-19 pandemic **Legend:** At the first two study visits, prior SARS-CoV-2 infection was detected by the presence of anti-S1-RBD-SARS-CoV-2 antibodies (orange). After the rollout of COVID-19-vaccines in early 2021, we assessed the presence of anti-NC-SARS-CoV-2 antibodies to differentiate between SARS-CoV-2-infection and COVID-19-vaccination related immune responses.

**Figure 2.**
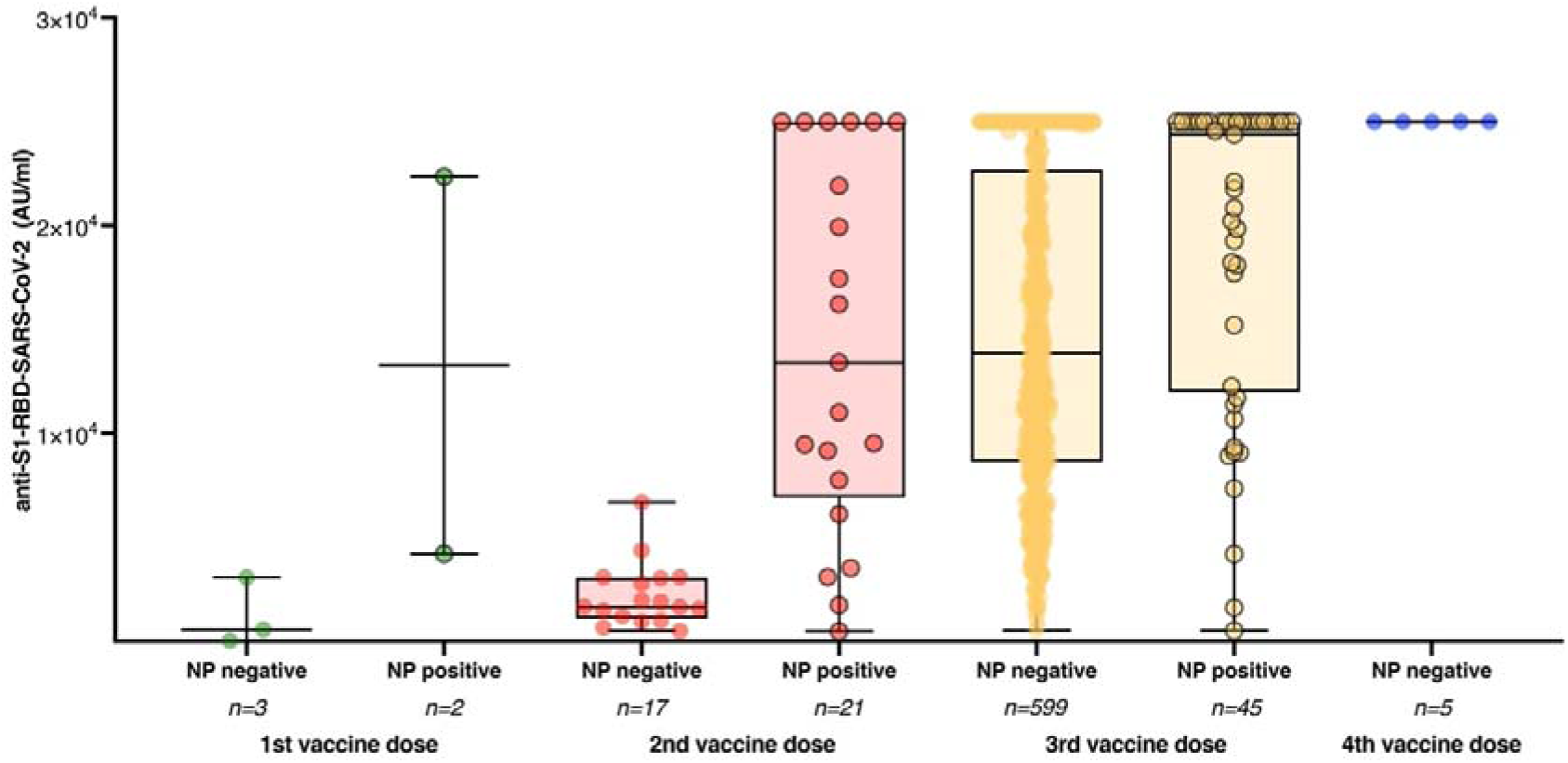
Anti-S1-RBD-SARS-CoV-2 antibody titers based on infection status and different vaccination regimens **Legend:** Anti-S1-RBD-SARS-CoV-2 antibody titers among study participants with (NP positive; dots with black edging) and without (NP negative; dots without black edging) detectable anti-NC-SARS-CoV-2 based on their vaccination status.

**Figure 3.**
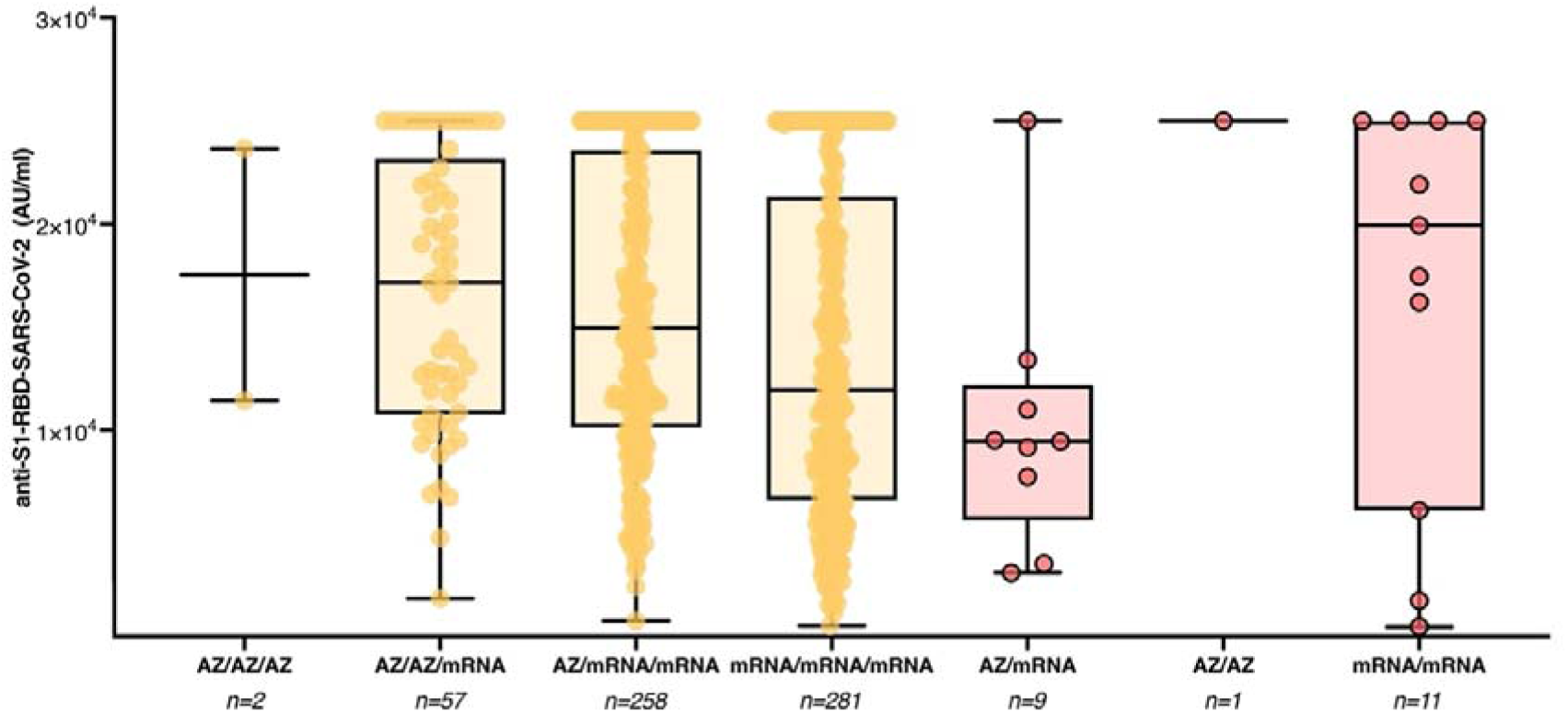
Anti-S1-RBD-SARS-CoV-2 antibody titers among study participants with three separate spike antigen exposures by COVID-19 booster vaccination or SARS-CoV-2 infection **Legend:** Anti-S1-RBD-SARS-CoV-2 antibody titers among study participants with three vaccine doses and no detectable anti-NC-SARS-CoV-2 antibodies (yellow) or two vaccine doses with and detectable anti-NC-SARS-CoV-2 antibodies (red).

## Discussion

We describe the SARS-CoV-2 infection and COVID-19 vaccination-induced immunity of 697 hospital workers at the University Medical Center Hamburg-Eppendorf between January 17 and 31, 2022. One of our key findings is that the prevalence of anti-NC-SARS-CoV-2 antibodies, indicating prior infection, was 9.8% at the current study visit, representing a doubling of the SARS-CoV-2 infection rate since the last study visit in May 2021. Of note, the official number of recovered COVID-19 patients in the city-state of Hamburg was 262’907 by the end of January 2022, which represents 14.2% of all inhabitants, assuming that no one had been infected more than once. While certainly some inhabitants were infected more than once, further unidentified or unreported COVID-19 cases must be assumed. In summary, the incidence rate among hospital workers in our study was considerably lower than in the general population, confirming that established infection control interventions remain effective in preventing nosocomial SARS-CoV-2 transmissions when the delta and omicron variants are the predominant strains. From our data, we conclude that a total of three spike antigen exposures by booster COVID-19 vaccination or any combination of SARS-CoV-2 infection with two vaccinations are necessary to achieve high antibody levels reliably. This supports current vaccination recommendations and is in line with previous studies demonstrating that triple-vaccinated naive individuals reach comparable levels of neutralization capacity against the omicron variant as vaccinated convalescents and twice-vaccinated individuals contracting break-through infections (Wratil et al., 2022). Likewise, efficacy studies showed that protection against SARS-CoV-2 infection in twice vaccinated individuals considerably wanes after six months, whereas convalescents boosted with two vaccine doses and triple-vaccinated individuals are protected for 12 months (Barda et al., 2021; Hall et al., 2022). Also, previous studies have shown that a single SARS-CoV-2 infection does not provide the same neutralization capacity as a combination of infection and vaccination (Wratil et al., 2022). Our findings are in line with previous investigations, which have shown that all licensed vaccines given as a homologous or heterologous third booster dose are effective in enhancing neutralizing antibody and cellular immune responses, notwithstanding which vaccines had been received in the initial vaccine course (Munro et al., 2021). In our study, the administration of a booster vaccine resulted in high antibody levels in the vast majority of participants regardless of the primary vaccination regimen, with a more than 7-fold increase between the second and the third vaccine dose. While among individuals with three vaccine doses, those with heterologous vaccination regimen had higher anti-S1-RBD-SARS-CoV-2 antibody levels among those compared to those who received three doses of an mRNA vaccine, multiple linear regression analysis revealed that only time since the booster vaccination and presence of anti-NC-SARS-CoV-2 antibodies, but not age, sex, and vaccination regimen significant predictors of anti-S1-RBD-SARS-CoV-2 antibody titers. Our study is subject to limitations. We did not recruit a strictly representative sample of hospital employees at our institution, which may limit the generalizability of results. Since employees who have been vaccinated may be more likely to participate in our study, we are not able to reliably assess the COVID-19 vaccination uptake among hospital workers at our institution. Employees with current SARS-CoV-2 infections may not have been able to participate in the current study visit, so we may underestimate the infection rate in our cohort to some degree. While 1253 and 872 hospital workers respectively participated in the last follow-up visits in June 2020 and May 2021, only 697 individuals participated at the current study visit. As in any real-world observational study, the participants were not randomly assigned to different COVID-19 vaccination regimens, and confounding may be expected due to dissimilarities in the different subgroups and differences in the time intervals between the respective vaccine doses and between the last vaccination and the current study visit. Also, given the limited sample size since we subsumed the mRNA vaccines BNT162b2 and mRNA-1273 to one single category and are therefore not able to assess potential differences in immunogenicity amongst different vaccination regimens with one or both of those vaccines. An additional limitation is that we quantitatively assessed the concentration of anti-SARS-CoV-2 antibodies but not their neutralizing capacity. While neutralizing antibody levels are highly predictive of immune protection from symptomatic SARS-CoV-2 infection (Khoury et al., 2021), the sheer concentration of antigen-specific antibodies may not be a reliable surrogate. Since the beginning of the pandemic, a combination of non-pharmaceutical interventions like social distancing, masking, testing, and contact tracing and the rollout of COVID-19 vaccines have been successful in mitigating the spread of SARS-CoV-2, especially among hospital employees and vulnerable patients. As the pandemic evolves, prospective longitudinal studies will be needed to investigate the duration of immune protection from circulating SARS-CoV-2 variants after COVID-19 vaccination, assess the necessity for novel omicron-targeted vaccines, and determine the timing for further booster vaccinations.

## Conclusion

The low prevalence of anti-NC-SARS-CoV-2 antibodies in our study cohort indicates the persistent effectiveness of established infection control interventions in preventing nosocomial transmission of delta and omicron SARS-CoV-2 variants. Our study further indicates that three exposures to the SARS-CoV-2 spike protein by either infection or vaccination are necessary to elicit particularly strong humoral immune responses, which supports the current COVID-19 vaccination recommendations.

## Data Availability

All data produced in the present work are contained in the manuscript

## Acknowledgments

We thank Valentina Davydov, Sabrina Kreß, Jennifer Wigger, Leon Cords and Maximilian Knapp for excellent technical assistance. We thank all study participants and departments of the University Medical Center Hamburg-Eppendorf for their active participation in the study.

---

**Supplemental Figure 1.**
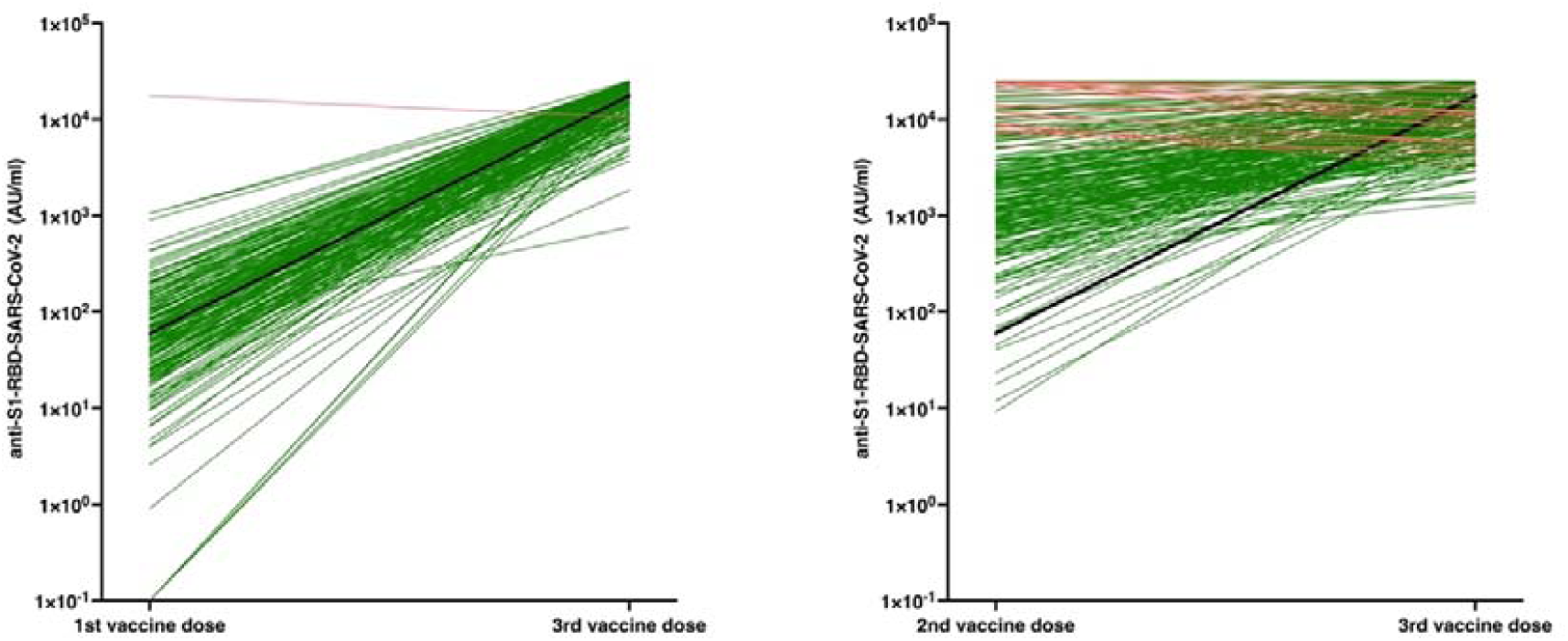
Kinetics of anti-S1-RBD-SARS-CoV-2 antibody titers between May 2021 and the current study visit **Legend:** Anti-S1-RBD-SARS-CoV-2 antibody titers are shown for study participants without anti-NC-SARS-CoV-2 antibodies indicating prior SARS-CoV-2 infection between the last study visit in May 2021 and the current study visit. The left panel shows the antibody titers for those who had received one vaccine dose in May 2021 and three vaccine doses by now. The right panel shows antibody titers for those who had received two vaccine dose in May 2021 and three vaccine doses by now. Increasing titers are portrayed as green lines, decreasing titers are portrayed as red lines. The fat black line connects the median titers of the two study visits.

**Supplemental Table 1.**

|  | **anti-NC negative (n=629)** | **anti-NC positive (n=68)** |
| --- | --- | --- |
| **Sex** |  |  |
| Male, n(%) | 131 (20.8) | 12 (17.6) |
| Female, n(%) | 491 (78.1) | 56 (82.4) |
| Diverse, n(%) | 7 (1.1) | 0 |
| **Age** |  |  |
| Median | 40 | 35 |
| IQR | 31; 51 | 28; 48 |
| **Profession** |  |  |
| Nurse, n(%) | 234 (37.2) | 27 (39.7) |
| Physician, n(%) | 130 (20.7) | 10 (14.7) |
| Medical Technician, n(%) | 100 (15.9) | 12 (17.6) |
| Other healthcare worker, n(%) | 75 (11.9) | 10 (14.7) |
| Non-healthcare worker, n(%) | 90 (14.3) | 9 (13.2) |

## References

Barda, N., Dagan, N., Cohen, C., Hernán, M.A., Lipsitch, M., Kohane, I.S., Reis, B.Y., Balicer, R.D., 2021. Effectiveness of a third dose of the BNT162b2 mRNA COVID-19 vaccine for preventing severe outcomes in Israel: an observational study. Lancet 398, 2093–2100.

Barros-Martins, J., Hammerschmidt, S.I., Cossmann, A., Odak, I., Stankov, M.V., Morillas Ramos, G., Dopfer-Jablonka, A., Heidemann, A., Ritter, C., Friedrichsen, M., Schultze-Florey, C., Ravens, I., Willenzon, S., Bubke, A., Ristenpart, J., Janssen, A., Ssebyatika, G., Bernhardt, G., Münch, J., Hoffmann, M., Pöhlmann, S., Krey, T., Bošnjak, B., Förster, R., Behrens, G.M.N., 2021. Immune responses against SARS-CoV-2 variants after heterologous and homologous ChAdOx1 nCoV-19/BNT162b2 vaccination. Nat Med 27, 1525–1529.

Borobia, A.M., Carcas, A.J., Pérez-Olmeda, M., Castaño, L., Bertran, M.J., García-Pérez, J., Campins, M., Portolés, A., González-Pérez, M., García Morales, M.T., Arana-Arri, E., Aldea, M., Díez-Fuertes, F., Fuentes, I., Ascaso, A., Lora, D., Imaz-Ayo, N., Barón-Mira, L.E., Agustí, A., Pérez-Ingidua, C., Gómez de la Cámara, A., Arribas, J.R., Ochando, J., Alcamí, J., Belda-Iniesta, C., Frías, J., 2021. Immunogenicity and reactogenicity of BNT162b2 booster in ChAdOx1-S-primed participants (CombiVacS): a multicentre, open-label, randomised, controlled, phase 2 trial. Lancet 398, 121–130.

Brehm, T.T., Schwinge, D., Lampalzer, S., Schlicker, V., Küchen, J., Thompson, M., Ullrich, F., Huber, S., Schmiedel, S., Addo, M.M., Lütgehetmann, M., Knobloch, J.K., Schulze Zur Wiesch, J., Lohse, A.W., 2021a. Seroprevalence of SARS-CoV-2 antibodies among hospital workers in a German tertiary care center: A sequential follow-up study. Int J Hyg Environ Health 232, 113671.

Brehm, T.T., Thompson, M., Ullrich, F., Schwinge, D., Addo, M.M., Spier, A., Knobloch, J.K., Aepfelbacher, M., Lohse, A.W., Lütgehetmann, M., Schulze Zur Wiesch, J., 2021b. Low SARS-CoV-2 infection rates and high vaccine-induced immunity among German healthcare workers at the end of the third wave of the COVID-19 pandemic. Int J Hyg Environ Health 238, 113851.

Hall, V., Foulkes, S., Insalata, F., Kirwan, P., Saei, A., Atti, A., Wellington, E., Khawam, J., Munro, K., Cole, M., Tranquillini, C., Taylor-Kerr, A., Hettiarachchi, N., Calbraith, D., Sajedi, N., Milligan, I., Themistocleous, Y., Corrigan, D., Cromey, L., Price, L., Stewart, S., de Lacy, E., Norman, C., Linley, E., Otter, A.D., Semper, A., Hewson, J., D’Arcangelo, S., Chand, M., Brown, C.S., Brooks, T., Islam, J., Charlett, A., Hopkins, S., 2022. Protection against SARS-CoV-2 after Covid-19 Vaccination and Previous Infection. N Engl J Med.

Karim, S.S.A., Karim, Q.A., 2021. Omicron SARS-CoV-2 variant: a new chapter in the COVID-19 pandemic. Lancet (London, England) 398, 2126–2128.

Khoury, D.S., Cromer, D., Reynaldi, A., Schlub, T.E., Wheatley, A.K., Juno, J.A., Subbarao, K., Kent, S.J., Triccas, J.A., Davenport, M.P., 2021. Neutralizing antibody levels are highly predictive of immune protection from symptomatic SARS-CoV-2 infection. Nat Med 27, 1205–1211.

Munro, A.P.S., Janani, L., Cornelius, V., Aley, P.K., Babbage, G., Baxter, D., Bula, M., Cathie, K., Chatterjee, K., Dodd, K., Enever, Y., Gokani, K., Goodman, A.L., Green, C.A., Harndahl, L., Haughney, J., Hicks, A., van der Klaauw, A.A., Kwok, J., Lambe, T., Libri, V., Llewelyn, M.J., McGregor, A.C., Minassian, A.M., Moore, P., Mughal, M., Mujadidi, Y.F., Murira, J., Osanlou, O., Osanlou, R., Owens, D.R., Pacurar, M., Palfreeman, A., Pan, D., Rampling, T., Regan, K., Saich, S., Salkeld, J., Saralaya, D., Sharma, S., Sheridan, R., Sturdy, A., Thomson, E.C., Todd, S., Twelves, C., Read, R.C., Charlton, S., Hallis, B., Ramsay, M., Andrews, N., Nguyen-Van-Tam, J.S., Snape, M.D., Liu, X., Faust, S.N., 2021. Safety and immunogenicity of seven COVID-19 vaccines as a third dose (booster) following two doses of ChAdOx1 nCov-19 or BNT162b2 in the UK (COV-BOOST): a blinded, multicentre, randomised, controlled, phase 2 trial. Lancet 398, 2258–2276.

Pozzetto, B., Legros, V., Djebali, S., Barateau, V., Guibert, N., Villard, M., Peyrot, L., Allatif, O., Fassier, J.B., Massardier-Pilonchéry, A., Brengel-Pesce, K., Yaugel-Novoa, M., Denolly, S., Boson, B., Bourlet, T., Bal, A., Valette, M., Andrieu, T., Lina, B., Cosset, F.L., Paul, S., Defrance, T., Marvel, J., Walzer, T., Trouillet-Assant, S., 2021. Immunogenicity and efficacy of □□□□□□□□□heterologous ChAdOx1-BNT162b2 vaccination. Nature 600, 701–706.

RKI 2022. https://impfdashboard.de (accessed 27.02.2022)

RKI 2021a. https://www.rki.de/DE/Content/Infekt/EpidBull/Archiv/2021/Ausgaben/43_21.pdf?blob=publicationFile (accessed 27.02.2022)

RKI 2021b. https://www.rki.de/DE/Content/Infekt/EpidBull/Archiv/2021/Ausgaben/48_21.pdf?blob=publicationFile (accessed 27.02.2022)

RKI 2021c. https://www.rki.de/DE/Content/Infekt/EpidBull/Archiv/2021/Ausgaben/27_21.pdf?blob=publicationFile (accessed 27.02.2022)

Roche 2022a. https://diagnostics.roche.com/global/en/products/params/elecsys-anti-sars-cov-2-s.html. (accessed 27.02.2022)

Roche 2022b. https://diagnostics.roche.com/global/en/products/params/elecsys-anti-sars-cov-2.html. (accessed 27.02.2022)

Wratil, P.R., Stern, M., Priller, A., Willmann, A., Almanzar, G., Vogel, E., Feuerherd, M., Cheng, C.C., Yazici, S., Christa, C., Jeske, S., Lupoli, G., Vogt, T., Albanese, M., Mejías-Pérez, E., Bauernfried, S., Graf, N., Mijocevic, H., Vu, M., Tinnefeld, K., Wettengel, J., Hoffmann, D., Muenchhoff, M., Daechert, C., Mairhofer, H., Krebs, S., Fingerle, V., Graf, A., Steininger, P., Blum, H., Hornung, V., Liebl, B., Überla, K., Prelog, M., Knolle, P., Keppler, O.T., Protzer, U., 2022. Three exposures to the spike protein of SARS-CoV-2 by either infection or vaccination elicit superior neutralizing immunity to all variants of concern. Nat Med.

